# Global associations of macronutrient supply and asthma disease burden

**DOI:** 10.1101/2023.12.18.23300109

**Authors:** Duan Ni, Alistair M. Senior, David Raubenheimer, Stephen J. Simpson, Laurence Macia, Ralph Nanan

## Abstract

**Objectives:** Global age-standardized prevalence of asthma has decreased over time, in parallel with which, the gross domestic product (GDP) per capita and nutritional landscapes also changed at a global level. Both socioeconomic status and nutritional factors are critical confounder for asthma, but most studies so far neglected to interrogate their correlations and interactions comprehensively. Hence, we aim to systematically investigate the relationship between nutrient supply, a good proxy of food environment, socioeconomic status and asthma disease burden at a global level over time.

**Methods:** Asthma disease burden, macronutrient (protein, carbohydrate and fat) supply and GDP data covering more than 150 countries around the globe from 1990 to 2018 was collated. Various multi-response generalized additive mixed models (GAMMs) were used to analyze the effects of macronutrient supplies and GDP over time on asthma disease burden.

**Results:** A model considering the interactions between macronutrient supplies and GDP, with an additive effect of time was favoured. Modelling results showed that carbohydrate supply was most strongly associated with increase of asthma disease burden, while fat supply had an opposite effect, and protein supply conferred less influences.

**Conclusions:** Globally, carbohydrate supply seems to play a driving role for asthma disease burden while fat supply might be the opposite. This is supported by previous studies about the amelioration of established asthma by ketogenic diet and might be linked to the diet quality. Further in-depth studies are warranted, which will be critical for future clinical research and practice and public health intervention.

## Main

We read with interest the work by Shin et al. ^1^, investigating the global disease burden of allergic disorders. They showed that the age-standardized prevalence of asthma has decreased over time on a global level.

Following a similar theme, we curated asthma disease burden, macronutrient (protein, carbohydrate and fat) supply and gross domestic product (GDP) data around the globe (Supplementary Methods). Our analysis found that in parallel to changes in asthma disease burden, GDP per capita ^2^ has increased, and the global nutritional landscape has also changed (Figure 1A-B). Both socioeconomic status and nutritional factors, are emerging critical confounders for asthma ^1^. However, so far, most studies have neglected to consider their correlations and interactions, including among nutrients within diets (Figure 1C-E). They instead focused on individual parameters alone, like specific diets or foods ^3^. Here, we adopted a recently published approach ^2^ to systematically interrogate the relationship between nutrient supply, a good proxy of food environment, socioeconomic status and asthma disease burden at a global scale over time. We focused on macronutrient supplies and their interactions, considering their important associations with many facets of health.

**Figure 1.**
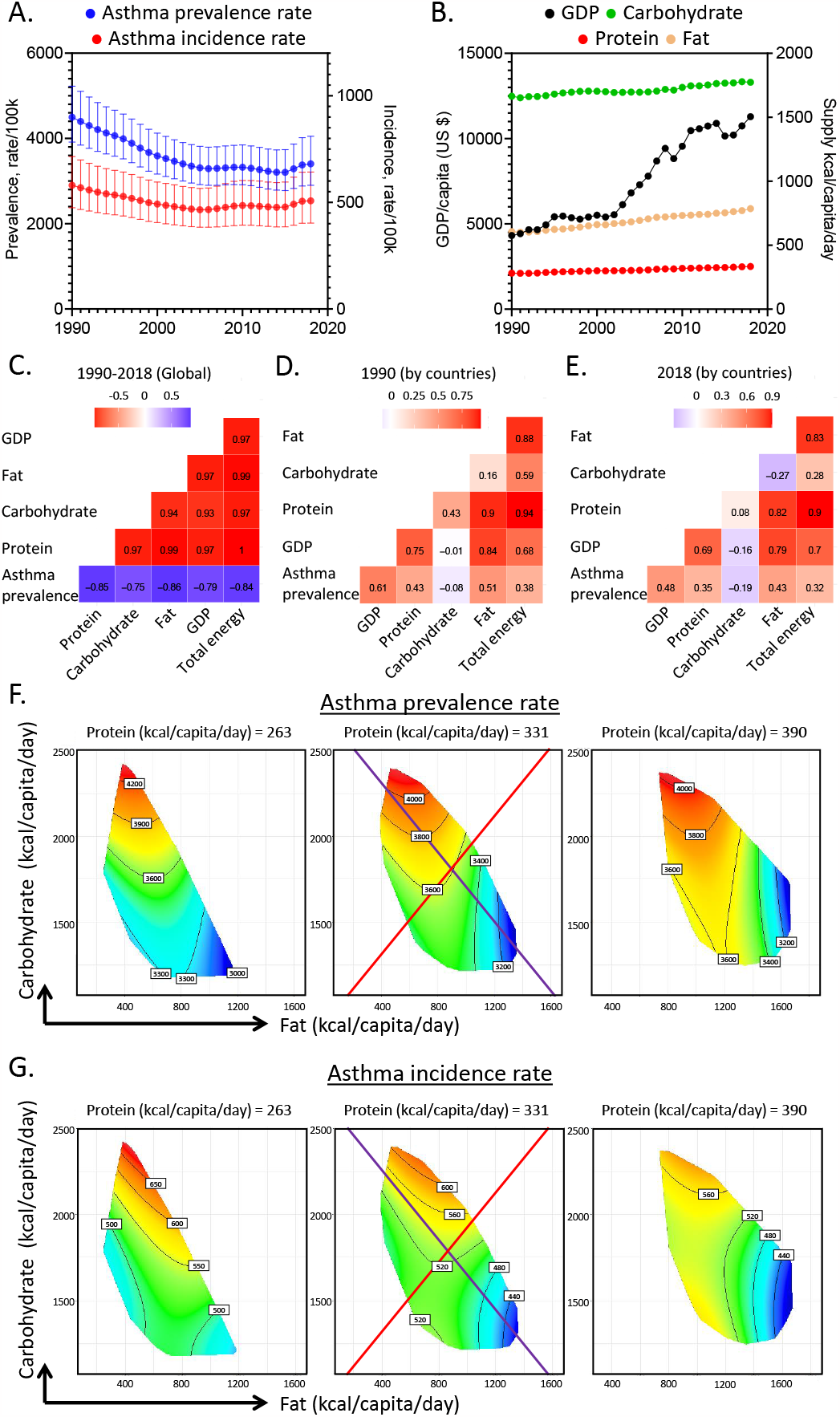
Association of global macronutrient supply and asthma disease burden. **A**. Age-standardized asthma prevalence (blue) and incidence rate (red) of both sexes as functions of year. **B**. Global GDP per capita (in US dollars, black) and supplies of carbohydrate (green), protein (red) and fat (brown) as functions of year. **C-E**. Correlations among variables for global data from 1990-2018 (**C**) and for different countries in 1990 (**D**) and 2018 (**E**). Correlation coefficients are shown. **F-G**. Predicted effects of macronutrient supply on age-standardized asthma prevalence rate (**F**) and incidence rate (**G**) of both sexes (See Supplementary Information for statistics and interpretation).

Various multi-response generalized additive mixed models (GAMMs) were used to analyze the effects on asthma disease burden from macronutrient supplies and GDP over time (Supplementary Methods). In all analyses, a model considering the interactions of macronutrient supply and GDP, and an additive effect of time was favoured (Supplementary Table 1, 3). This suggested combinatory effects of macronutrients on asthma and the impacts of socioeconomic changes.

The most recent year with all data available, 2018, is shown as a representative (Figure 1F-G, Supplementary Table 1-2). In brief, predicted association between asthma prevalence and macronutrient supplies were visualized using response surfaces on macronutrient supply plots. We focused on the fat (*x*-axis) and carbohydrate (*y*-axis) supplies while holding protein at 25%, 50% and 75% quantiles of global supply. Across the response surfaces, red indicated higher asthma disease burden, while blue indicated lower.

In our modelling, carbohydrate supply was most strongly associated with increases of asthma prevalence rates, while fat supply had the opposite effects (Figure 1F). This is reflected by the purple isocaloric line, along which the total energy from macronutrients remained constant but increasing fat:carbohydrate ratio decreased the allergy prevalence. Protein supply conferred less influences. Similar patterns were found for asthma incidences (Figure 1G, Supplementary Table 3-4) independent of gender (Supplementary Figure 1-2) and were not confounded by the total macronutrient energy supply, as changing total energy while holding fat:carbohydrate ratio constant (red radial in figures) minimally impacted asthma disease burden.

This represents the first study to link asthma to global food environment. Our results imply a driving role of carbohydrate supply for the asthma disease burden, after adjusting for the plausible interactions between macronutrients, total energy supply and socioeconomic status.

Interestingly, previous studies found that ketogenic diets, low in carbohydrates, might ameliorate established asthma ^4^, supporting our findings that ketogenic-like food environments are associated with lower asthma disease burden. Although further in-depth investigations are needed, diet quality might be an intriguing explanation for the positive association between carbohydrate supply and asthma. For example, high- and ultra-processed foods low in fibre have been found to be related to asthma development ^3,5^, while we have previously shown that dietary fibre exhibited a strong immune regulatory influence, protecting against asthma ^6^.

Hence, future studies in more depth are warranted to investigate the associations between macronutrient supply and asthma, including also other related socioeconomic and environmental factors. Such studies will be critical for guidance to future clinical research and practice and public health interventions.

## Data Availability

All data produced in the present study are available upon reasonable request to the authors

## Author contributions

*Concept and design:* Duan Ni, and Ralph Nanan

*Acquisition, analysis and interpretation of data:* Duan Ni, Alistair M. Senior, Ralph Nanan

*Drafting of the manuscript:* Duan Ni, and Ralph Nanan

*Critical revision of the manuscript for important intellectual content:* All authors

## Acknowledgements

This project is supported by the Norman Ernest Bequest Fund.

