## Supplementary Information for "Global associations of macronutrient supply and asthma disease burden"

**Interpretation of modelling surfaces**

Details for modelling surface interpretation is adapted from our previous publications^1,2^. In brief, using RStudio (v4.2.2.), asthma disease burden data were analyzed with generalized additive mixed models (GAMMs) prediction^3,4^ and plotted as response surfaces based on nutrient axes for carbohydrate and fat, while holding protein supply at a range of constants (taken to be 25%, 50% (median) and 75% quantiles of the global protein supply of different countries). The surfaces depict the relationship between asthma disease burden and the supply of protein, carbohydrate and fat (kcal/capita/day). The protein supply increases from left to right across the plots, while the supply of fat and carbohydrate changes along X and Y axes within each plot. Within surfaces, red implies higher values while blue indicates lower ones. The modelling values remain unchanged along the black contour lines on the surfaces and the numbers on them denote the magnitude of the parameters. The purple line is an isocaloric line, along which the total energy supply from macronutrients is unchanged but the ratio of carbohydrate and fat supply differs. The red line is a food rail. Carbohydrate:fat ratio is fixed along the line but the total energy supply from macronutrients is altered.

Statistics for the modelling analysis is provided in Supplementary Tables. When the modelling analysis is significant, the effect of macronutrient supply and their combinations on the modelling values (i.e., asthma disease burden) can be deduced from the modelling surfaces.

**Supplementary Methods**

*Data collection and processing*

Asthma disease burden data was downloaded from Global Burden of Disease Study 2019 (GBD 2019). As previously described^1^, macronutrient supply data was extracted from the Food and Agriculture Organization Corporate Statistical Database (FAOSTAT, [www.fao.org/faostat/en/#home](http://www.fao.org/faostat/en/#home)), while the gross domestic product (GDP) per capita data was collected from the Maddison project^5^. Since GBD reported data from 1990 to 2018, analyses were thus based on this period of time. After collating disease, nutrient and GDP data and filtering for countries or timepoints without data record, data spanning 1990-2018 across 159 countries were left for further analysis with R.

*Generalized additive mixed models (GAMMs)*

Details of GAMM analysis was described in^1^. In brief, GAMMs^3,4^ were deployed to assess changes in asthma disease burden over time and the effects from macronutrient supply and GDP. GAMMs take into consideration nonlinear terms as nonparametric smoothed functions, which are often a form of spline, and provide a flexible manner to estimate nonlinear associations. All models were implemented with the *mgcv* package and its “gam” function^3,4^. All models considered the country that the data were based on as a random effect. In all models, the gamma parameter, reflecting the degree to which the modelled effects are smoothed was defined as log(*n*)/2 (*n* is the number of combinations for country and year with available data). A Gaussian family with log-link function was used for modelling.

We compared a number of different predictor variables and in different combinations alongside a null model where only the random effect from the country is considered. Other models took into account all combinations of the individual, additive and interactions among macronutrient supply, year and GDP data. Macronutrient supply was modelled as a three dimensional spline, while year and GDP were modelled as single dimensional cubic-regression splines using the “s()” function in *mgcv* package. “te()” function from *mgcv* package was used to model the interactions between smooth terms existing on different scales like macronutrient supply and year based on tensor product smoothers.

Modelling results were compared based on their Akaike information criterions (AICs) and the model with the lowest AIC was selected^6^.

Codes for the analysis can be found at GitHub, <https://github.com/Nidane/Asthma-NutrientSupply>.

**Supplementary Tables**

Supplementary Tables 1-4 are GAMMs estimates. For parametric terms the model estimates and associated standard errors (SE) and test statistics are presented. For non-parametric smooth terms, the estimated and reference degrees of freedom (reflected by edf, sumEDF, Ref.df) are shown as well as their test statistics. Smooth terms were fitter with either the standard smooth function “s()” or a tensor product smooth “te()” in the *mgcv* package. Tensor product smoothing was utilized where terms exist on different units (for example, nutrient supply and year). Country was included as a random effect using the smooth function “s()”. The model family and implemented link function is stated.

**Supplementary Table 1.** Relative fit of generalized additive mixed models (GAMMs) testing the predictors for age-standardized asthma prevalence rate in both sexes. Gamma is the degrees of freedom inflation factor. Dev means the deviance explained. AIC = Akaike information criterion. GDP = gross domestic product. Delta is the differences between AICs of models and the minimum AIC. sumEDF reflects the degrees of freedom of the models. Macronutrient supply was modelled as a three dimensional thin-plate spline. (related to Figure 1F)

| **GAM** | **Gamma** | **Dev** | **AIC** | **Delta** | **Weights** | **sumEDF** | **Formula** |
| --- | --- | --- | --- | --- | --- | --- | --- |
| 1 | 3.8554 | 92.13 | 70355.5 | 4467.2 | 0 | 154.815 | 1 + s(Country, bs="re") |
| 2 | 3.8554 | 93.45 | 69565.1 | 3676.8 | 0 | 167.432 | s(protein.kcal, carb.kcal, fat.kcal, k=k_nut) + s(Country, bs="re") |
| 3 | 3.8554 | 96.32 | 66980.7 | 1092.4 | 6.27E-238 | 161.693 | s(Year, k=10, bs="cr") + s(Country, bs="re") |
| 4 | 3.8554 | 94.7 | 68619.4 | 2731.1 | 0 | 166.544 | s(GDP, k=10, bs="cr") + s(Country, bs="re") |
| 5 | 3.8554 | 96.52 | 66749.2 | 860.95 | 1.11E-187 | 172.529 | s(protein.kcal, carb.kcal, fat.kcal, k=k_nut) + s(Year, k=10, bs="cr") + s(Country, bs="re") |
| 6 | 3.8554 | 95.21 | 68184.8 | 2296.5 | 0 | 176.641 | s(protein.kcal, carb.kcal, fat.kcal, k=k_nut) + s(GDP, k=10, bs="cr") + s(Country, bs="re") |
| 7 | 3.8554 | 96.45 | 66826.1 | 937.83 | 2.25E-204 | 168.17 | s(Year, k=10, bs="cr") + s(GDP, k=10, bs="cr") + s(Country, bs="re") |
| 8 | 3.8554 | 96.64 | 66636.5 | 748.16 | 3.46E-163 | 191.222 | te(protein.kcal, carb.kcal, fat.kcal, Year, bs=c("tp", "cr"), d=c(3,1), k=c(k_nut, 7)) + s(Country, bs="re") |
| 9 | 3.8554 | 96.33 | 67039.9 | 1151.6 | 8.47E-251 | 199.688 | te(protein.kcal, carb.kcal, fat.kcal, GDP, bs=c("tp", "cr"), d=c(3,1), k=c(k_nut, 7)) + s(Country, bs="re") |
| 10 | 3.8554 | 96.79 | 66414.8 | 526.54 | 4.60E-115 | 188.532 | te(Year, GDP, k=10) + s(Country, bs="re") |
| 11 | 3.8554 | 96.74 | 66508.6 | 620.3 | 2.01E-135 | 200.283 | te(protein.kcal, carb.kcal, fat.kcal, Year, bs=c("tp", "cr"), d=c(3,1), k=c(k_nut, 7)) + s(GDP, k=10, bs="cr") + s(Country, bs="re") |
| **12** | **3.8554** | **97.16** | **65888.3** | **0** | **1** | **196.77** | **te(protein.kcal, carb.kcal, fat.kcal, GDP, bs=c("tp", "cr"), d=c(3,1), k=c(k_nut, 7)) + s(Year, k=10, bs="cr") + s(Country, bs="re")** |
| 13 | 3.8554 | 96.96 | 66198.5 | 310.21 | 4.35E-68 | 199.798 | te(Year, GDP, k=10) + s(protein.kcal, carb.kcal, fat.kcal, k=k_nut) + s(Country, bs="re") |

**Supplementary Table 2.** Estimated effects of macronutrient supply by time and GDP per capita on age-standardized asthma prevalence rate. Gaussian-GAMM, log-link function. (related to Figure 1F)

| **Parametric coefficients** | |  |  |  |
| --- | --- | --- | --- | --- |
|  | **Estimate** | **Std. Error** | **t value** | **Pr(>\|t\|)** |
| **(Intercept)** | 8.2765 | 0.04056 | 204.1 | <2e-16 |
| **Approximate significance of smooth terms** | | | |  |
|  | **edf** | **Ref.df** | **F** | **p-value** |
| **te(protein.kcal,carb.kcal,fat.kcal,GDP)** | 34.072 | 38.087 | 31.55 | <2e-16 |
| **s(Year)** | 5.144 | 6.239 | 246.53 | <2e-16 |
| **s(Country)** | 157.555 | 158 | 451.94 | <2e-16 |
| R-sq.(adj) = 0.97 Deviance explained = 97.2% | | | | |
| GCV = 1.9963e+05 Scale est. = 1.4363e+05 n = 4465 | | | | |

**Supplementary Table 3.** Relative fit of generalized additive mixed models (GAMMs) testing the predictors for age-standardized asthma incidence rate in both sexes. Gamma is the degrees of freedom inflation factor. Dev means the deviance explained. AIC = Akaike information criterion. GDP = gross domestic product. Delta is the differences between AICs of models and the minimum AIC. sumEDF reflects the degrees of freedom of the models. Macronutrient supply was modelled as a three dimensional thin-plate spline. (related to Figure 1G)

| **GAM** | **Gamma** | **Dev** | **AIC** | **Delta** | **Weights** | **sumEDF** | **Formula** |
| --- | --- | --- | --- | --- | --- | --- | --- |
| 1 | 3.8554 | 94.03 | 48074.2 | 3690.9 | 0 | 156.384 | 1 + s(Country, bs="re") |
| 2 | 3.8554 | 95.07 | 47248.6 | 2865.3 | 0 | 167.318 | s(protein.kcal, carb.kcal, fat.kcal, k=k_nut) + s(Country, bs="re") |
| 3 | 3.8554 | 96.05 | 46247.1 | 1863.9 | 0 | 161.01 | s(Year, k=10, bs="cr") + s(Country, bs="re") |
| 4 | 3.8554 | 94.99 | 47314.4 | 2931.1 | 0 | 166.368 | s(GDP, k=10, bs="cr") + s(Country, bs="re") |
| 5 | 3.8554 | 96.3 | 45975.8 | 1592.5 | 0 | 171.426 | s(protein.kcal, carb.kcal, fat.kcal, k=k_nut) + s(Year, k=10, bs="cr") + s(Country, bs="re") |
| 6 | 3.8554 | 95.46 | 46894 | 2510.8 | 0 | 175.92 | s(protein.kcal, carb.kcal, fat.kcal, k=k_nut) + s(GDP, k=10, bs="cr") + s(Country, bs="re") |
| 7 | 3.8554 | 96.16 | 46126 | 1742.8 | 0 | 168.804 | s(Year, k=10, bs="cr") + s(GDP, k=10, bs="cr") + s(Country, bs="re") |
| 8 | 3.8554 | 96.54 | 45694 | 1310.8 | 2.35E-285 | 185.31 | te(protein.kcal, carb.kcal, fat.kcal, Year, bs=c("tp", "cr"), d=c(3,1), k=c(k_nut, 7)) + s(Country, bs="re") |
| 9 | 3.8554 | 96.9 | 45284.8 | 901.53 | 1.72E-196 | 222.565 | te(protein.kcal, carb.kcal, fat.kcal, GDP, bs=c("tp", "cr"), d=c(3,1), k=c(k_nut, 7)) + s(Country, bs="re") |
| 10 | 3.8554 | 96.58 | 45650.9 | 1267.6 | 5.45E-276 | 187.693 | te(Year, GDP, k=10) + s(Country, bs="re") |
| 11 | 3.8554 | 96.61 | 45627.8 | 1244.6 | 5.55E-271 | 196.065 | te(protein.kcal, carb.kcal, fat.kcal, Year, bs=c("tp", "cr"), d=c(3,1), k=c(k_nut, 7)) + s(GDP, k=10, bs="cr") + s(Country, bs="re") |
| **12** | **3.8554** | **97.47** | **44383.3** | **0** | **1** | **226.63** | **te(protein.kcal, carb.kcal, fat.kcal, GDP, bs=c("tp", "cr"), d=c(3,1), k=c(k_nut, 7)) + s(Year, k=10, bs="cr") + s(Country, bs="re")** |
| 13 | 3.8554 | 96.74 | 45451.7 | 1068.4 | 9.89E-233 | 197.613 | te(Year, GDP, k=10) + s(protein.kcal, carb.kcal, fat.kcal, k=k_nut) + s(Country, bs="re") |

**Supplementary Table 4.** Estimated effects of macronutrient supply by time and GDP per capita on age-standardized asthma incidence rate. Gaussian-GAMM, log-link function. (related to Figure 1G)

| **Parametric coefficients** | |  |  |  |
| --- | --- | --- | --- | --- |
|  | **Estimate** | **Std. Error** | **t value** | **Pr(>\|t\|)** |
| **(Intercept)** | 6.31966 | 0.01529 | 413.3 | <2e-16 |
| **Approximate significance of smooth terms** | | | |  |
|  | **edf** | **Ref.df** | **F** | **p-value** |
| **te(protein.kcal,carb.kcal,fat.kcal,GDP)** | 65.665 | 68.310 | 34.32 | <2e-16 |
| **s(Year)** | 4.317 | 5.311 | 209.05 | <2e-16 |
| **s(Country)** | 156.650 | 158 | 663.41 | <2e-16 |
| R-sq.(adj) = 0.973 Deviance explained = 97.5% | | | | |
| GCV = 1698.8 Scale est. = 1155.5 n = 4465 | | | | |

**Supplementary Figures**

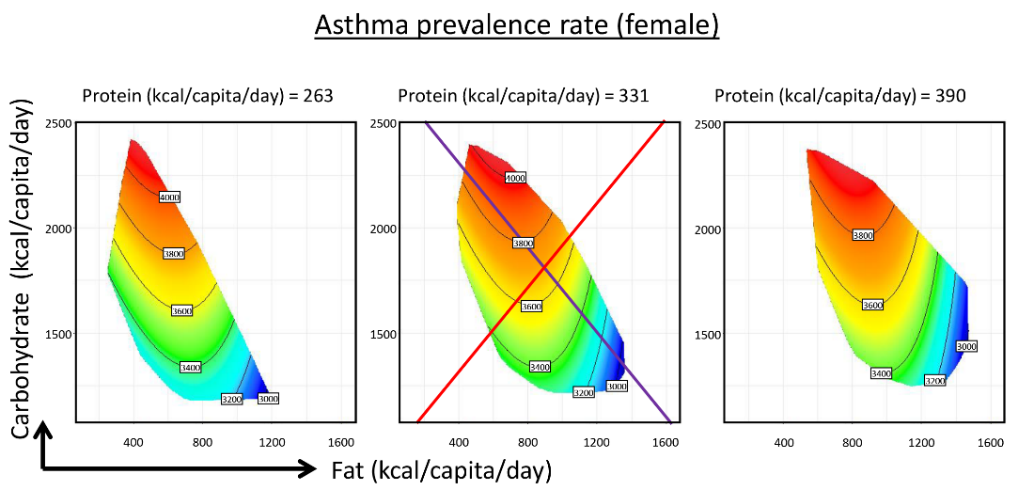

**Supplementary Figure 1.** Predicted effects of macronutrient supply on age-standardized asthma prevalence rate of female.

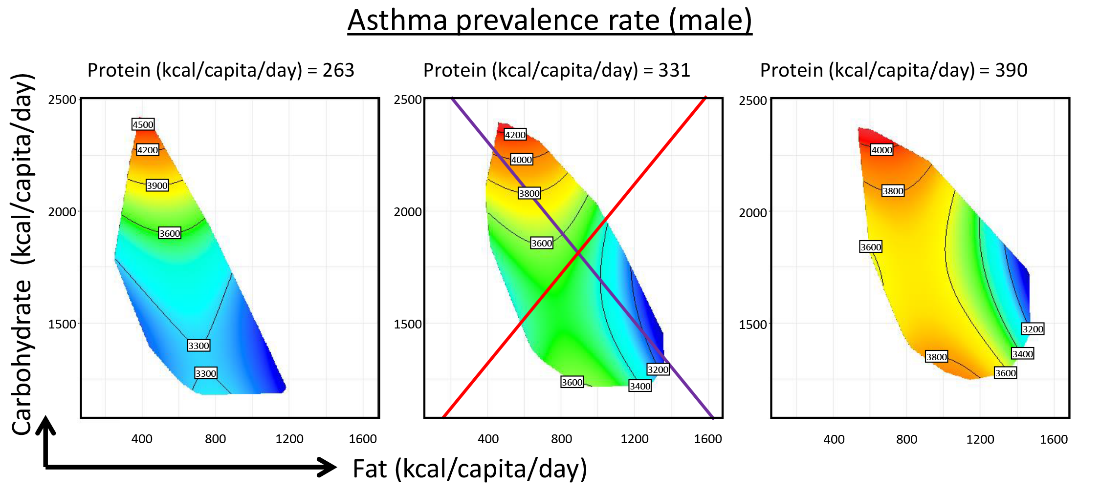

**Supplementary Figure 2.** Predicted effects of macronutrient supply on age-standardized asthma prevalence rate of male.
